# Paramagnetic Rim Lesions are Highly Specific for Multiple Sclerosis in Real-World Data

**DOI:** 10.1101/2024.08.14.24312000

**Authors:** Christopher C. Hemond, Sathish K. Dundamadappa, Mugdha Deshpande, Jonggyu Baek, Robert H. Brown, Carolina Ionete, Daniel S. Reich

**Affiliations:** Departments of Neurology; Radiology or; Population and Quantitative Health Sciences, University of Massachusetts Memorial Medical Center and University of Massachusetts Chan Medical School, Worcester, MA, USA; Translational Neuroradiology Section, National Institute of Neurological Disorders and Stroke, National Institutes of Health, Bethesda, MD, USA

## Abstract

**Background:** Paramagnetic rim lesions (PRL) are an emerging biomarker for multiple sclerosis (MS). In addition to associating with greater disease severity, PRL may be diagnostically supportive.

**Objective:** Our aim was to determine PRL specificity and sensitivity for discriminating MS from its diagnostic mimics using real-world clinical diagnostic and imaging data.

**Methods:** This is a retrospective, cross-sectional analysis of a longitudinal cohort of patients with prospectively collected observational data. Patients were included if they underwent neuroimmunological evaluation in our academic MS center, and had an available MRI scan from the same clinical 3T magnet that included a T2*-weighted sequence with susceptibility postprocessing (SWAN protocol, GE). SWAN-derived filtered phase maps and corresponding T2-FLAIR images were manually reviewed to determine PRL. PRL were categorized as “definite,” “probable,” or “possible” based on modified, recent consensus criteria. We hypothesized that PRL would convey a high specificity to discriminate MS from its MRI mimics.

**Results:** 580 patients were evaluated in total: 473 with MS, 57 with non-inflammatory neurological disease (NIND), and 50 with other inflammatory neurological disease (OIND). Identification of “definite” or “probable” PRL provided a specificity of 98% to discriminate MS from NIND and OIND; sensitivity was 36%. Interrater agreement was almost perfect for definite/probable identification at a subject level.

**Conclusions:** PRL convey high specificity for MS and can aid in the diagnostic evaluation. Modest sensitivity limits their use as single diagnostic indicators. Including lesions with lower confidence (“possible” PRL) rapidly erodes specificity and should be interpreted with caution given the potential harms associated with misdiagnosis.

## Introduction

Multiple sclerosis (MS) is a chronic inflammatory and degenerative disease of the central nervous system that benefits from early treatment with disease-modifying therapies ^1^. Real-world data suggest concerning rates of misdiagnosis^2^, a problem that highlights an ongoing need for MS biomarkers with high specificity. The paramagnetic rim lesion (PRL) is a candidate MRI marker in this regard and has been shown in a series of studies to be associated with high (≥95%) specificity for MS^3,4^. However, most studies assessing PRL have used research protocols and cohorts that limit generalizability of the findings. Moreover, PRL categorization is a qualitative endeavor, and the pursuit of perfect inter-rater agreement has been challenging^5^. Although recent NAIMS consensus criteria for determination of PRL will aid in standardization of PRL identification^6^, these metrics have yet to be systematically evaluated in the real world.

In this study, we aimed to determine the specificity and sensitivity of PRL through an analysis of clinical and MRI data from a single academic neuroimmunology clinic. These data reflect the wide variety of patients typically seen in clinical practice who are often referred for MS diagnostic consideration and management. We additionally aimed to create and evaluate a system for rating PRL confidence that could aid in guiding clinical relevance of lesion assessment features.

## Methods

### Consent to Participate

This study was reviewed and approved by the University of Massachusetts ethics board (IRB Protocols # H00016906 and 14143). Data collection, storage, and access were in accordance with the Health Insurance Portability and Accountability Act. All participants provided written informed consent.

### Study Design and Subjects

This is a retrospective analysis of prospectively collected data from patients at a single, large, academic neuroimmunology clinic; the study timeframe was March 2016 to August 2022. All subjects were participating in at least one of two ongoing prospective, longitudinal observational studies of neuroimmunological disorders. Inclusion criteria were: age less than 80 years, having been evaluated in outpatient neuroimmunology clinic for neurological symptoms, and having had at least one MRI performed on the same outpatient 3T MRI scanner with an “MS” protocol including susceptibility-sensitive imaging. Baseline was considered as the time of the first eligible MRI, with clinical measures calculated relative to that value.

Patient neurological disability ratings and disease-modifying therapy (DMT) use were procured by review of the patient electronic medical record and chosen based on proximity to the time of the baseline MRI. All EDSS scores and MS diagnoses/phenotypes extracted from the chart were rated by subspecialty-trained neuroimmunologists. The Multiple Sclerosis Severity Score (MSSS)^7^ and Age-Related MS Severity Score^8^ were calculated based on tabulated values when possible; MSSS was not calculated for disease durations of less than 6 months.

Patient diagnoses were extracted by chart review, using the latest information available. Diagnoses were made only by one of five expert neuroimmunologists, all of whom had subspecialty clinical training. All diagnoses for CIS and MS met 2017 McDonald^9^ criteria. Radiologically isolated syndromes (RIS) were defined based on the updated (2023) criteria.^10^ Other inflammatory neurological disease (OIND) diagnosis must meet criteria for one of the following: (1) neuromyelitis optica spectrum disorder (NMOSD), (2) myelin oligodendrocyte glycoprotein-associated disease (MOGAD), (3) proven or highly probable CNS infection or granulomatous disease, (4) T2-hyperintense lesions and/or inflammatory cerebrospinal fluid profiles with concomitant diagnosis of a systemic inflammatory/autoimmune disease felt by their clinician to be causative or associated with CNS injury. “Possible MS” was defined as nonspecific neurological symptoms in the presence of at least one white matter brain lesion fulfilling DIS candidate areas, but without meeting revised RIS 2023 criteria; these patients were subsumed under OIND. Non-inflammatory neurological disease (NIND) included all other non-MS patients not meeting OIND criteria.

### Image Acquisition

All images were acquired on a 3T MRI scanner (Signa Pioneer, General Electric Healthcare, Wisconsin, USA) using a 32-channel head and neck coil and a consistent acquisition protocol for MS patients. This standard protocol was implemented beginning in September 2016. There were no hardware upgrades that occurred during the observation period; 3 software updates occurred over the observation period of this study (PX25, PX26, and PX29), with two, minor additional upgrades within each version. The protocol included sagittal 3D T2-FLAIR (FLuid Attenuated Inversion Recovery): TR/TE/flip angle/echo train length=5400/maximum ∼133/90°/140, frequency/phase=256/224, slice thickness=0.8 at 50% resolution, reconstructed voxel size=0.49×0.49×0.80mm, FOV=250mm, scan duration=3:34. Most susceptibility-sensitive scans (93.3%) were obtained using the following protocol: 3D susceptibility-sensitive imaging (Susceptibility Weighted ANgiography, i.e. SWAN protocol): TR/TE/flip angle/echo train length=minimum ∼42ms/minimum ∼24ms/10°/6, frequency/phase=320/224, slice thickness/overlap=3.0mm/1.5mm, reconstructed voxel size=0.47×0.47×3.0mm, FOV=240mm, scan duration=2:27; after November 2021 the following parameters with changed (affecting 6.7% of scans): flip angle/echo train length=8°/10, frequency/phase=256/256, slice thickness/overlap=2.0mm/1.0mm, FOV=220mm. If gadolinium was administered, an additional sagittal 3D T1-weighted fast spin echo (CUBE, no acronym) scan was obtained approximately 10 minutes after the intravenous infusion of gadoterate meglumine with the following parameters: TR/TE/flip angle 435ms/16.1ms/90°, frequency/phase=256/256, slice thickness=0.6, reconstructed voxel size=0.49×0.49×0.60mm.

### Image Analysis

MRI data underwent reconstruction with the scanner’s manufacturer software and were exported in DICOM format. Images were converted to NIFTI format using dcm2niix software (version 1.0.20201102) ^11^. T1-weighted scans were resliced to a resolution of 0.80mm isotropic. Manufacturer-reconstructed Susceptibility-Weighted ANgiography (SWAN) filtered phase images, adhering to the “right hand” convention where paramagnetic substances appear dark, along with T2-FLAIR sequences, were aligned (using mutual information criteria and a rigid transformation with 6 degrees of freedom) with SPM software (available at https://www.fil.ion.ucl.ac.uk/spm/software/spm12/). FLAIR and filtered-phase SWAN images were examined side-by-side in the axial view by one or both raters (C.C.H., a neurologist with 6 years of post-training experience with PRL specifically, and S.K.D., a neuroradiologist with 17 years of post-training experience; 6 with PRL) using ITK-SNAP software ^12^, with both evaluators blinded to the clinical details during the assessment. All PRL were evaluated at a standardized contrast setting; intensity histograms for GE phase images ranged between -π and - π, scaled by a factor of ∼1000. Our evaluation widow was set between -300 and 300 with no adjustment of additional control points (linear mapping).

### PRL Determination

PRL were defined as FLAIR lesions with a colocalized paramagnetic rim as seen on the filtered-phase SWAN in the axial plane. Raters were free to visualize PRL in the (lower resolution) orthogonal planes at their discretion; in practice this occurred only rarely. The paramagnetic rim was required to be contiguous through at least 2/3 of the border (excepting ventricular and cortical margins) and must be visualized in at least 2 axial slices. The core of the PRL must be similarly isointense (with small variations accepted) to normal appearing white matter. These criteria are similar to the recent NAIMS consensus criteria ^6^, with the differences being that we did not immediately exclude lesions with complex or edge-like venous features. We additionally implemented a categorical “confidence” assessment system for raters based on judgements of lesion features that contributed uncertainty. These “flag” categories were defined as: (1) heterogeneity in core or rim; (2) incomplete paramagnetic border; (3) lesion within a confluent complex; (4) faint paramagnetic signal; (5) vascular complexity near lesion rim; (6) incomplete edge colocalization between paramagnetic rim and FLAIR lesion edge; (7) small size (< ∼5mm long-axis diameter) and (8) susceptibility artifacts (proximate to dental hardware or air/bone interfaces, or major dipole phase inversion). Milder dipole edge artifacts (phase inversion outside the paramagnetic rim) were not recorded. Raters marked their level of confidence based on the number and severity of these categories. Up to two of the most contributory conditions that reduced confidence were recorded. Because the severity of these confidence-reducing features was variable, we ultimately allowed rater judgement as the final arbitrating factor, akin to a real-world radiology setup.

Lesions that were part of confluent complexes were included for assessment. PRL could not be considered “definite” or “probable” if they did not share a significant edge portion with the FLAIR signal, however. All lesions were assessed for contrast-enhancement and excluded if they exhibited this property. Scans that did not include contrast were either (1) compared to a prior available scan to ensure lesion presence and chronicity or (2) compared to a subsequent susceptibility-sensitive scan to ensure PRL chronicity. Cases in which these conditions were not met (N=8) were few, but remained included in the analysis after chart review revealed that none of these patients were having new symptoms at the time of the scan, and the lesions in question did not exhibit any restricted diffusion or signs of edema that might be suggestive of active demyelination. In cases where PRL could not be confirmed at follow-up (and no available prior, N=2), patients had at least one definite/probable PRL that was already confirmed, such that this would not change patient category of ≥1 vs 0.

### Interrater Assessment

We randomly assessed 43 patients without confluent lesions to determine inter-rater agreement. No attempts were made to reconcile findings, and the analyzed results reflect the original rater only (N=465 rated by CH, N=115 rated by SD with 43 overlap). Cohen’s Kappa was used to determine interrater agreement measures as implemented in the R toolbox “IRR” at the patient-level (≥1 PRL with and between confidence categories). To coordinate interrater comparisons, T2-hyperintense lesion maps were segmented from T2-FLAIR images using the lesion prediction algorithm^13^ as implemented in the LST toolbox version 3.0.0 (www.statistical-modelling.de/lst) for SPM. A lesion index was created using the “cluster” function in FSL, to label individual lesions. Lesion segmentation failures (N=29) or scans with substantial confluent lesions (N=44) were assessed by a single rater. LST failures were later segmented using SAMSEG^14^ to obtain lesion number and volumes.

### Statistical Analysis

We assessed normality using a combination of skew/kurtosis metrics and histogram inspection. All statistical analyses were performed in R-studio (www.r-studio.com) version 4.02, with libraries including Tidyverse^15^, yardstick (sensitivity and specificity)^16^, and IRR (interrater agreement)^17^.

### Timed Analysis

One rater (CH) timed their analysis of randomly assessed patients and determined the number of seconds to (1) first PRL detection and (2) total count of PRL detected.

## Results

### Cohort description

A total of 580 patients met initial inclusion criteria. Of these, 473 were diagnosed with MS (relapsing = 375, progressive = 98), 57 with NIND, and 50 with OIND (N=8 clinically isolated syndrome and N=3 radiologically isolated syndrome were included in the relapsing category). A summary of these patient populations can be seen in **Table 1**. Relapsing MS patients were notably younger (45±12 years) compared to the progressive (58±9 years), NIND (50±14), and OIND (51±12) cohorts.

**Table 1:**
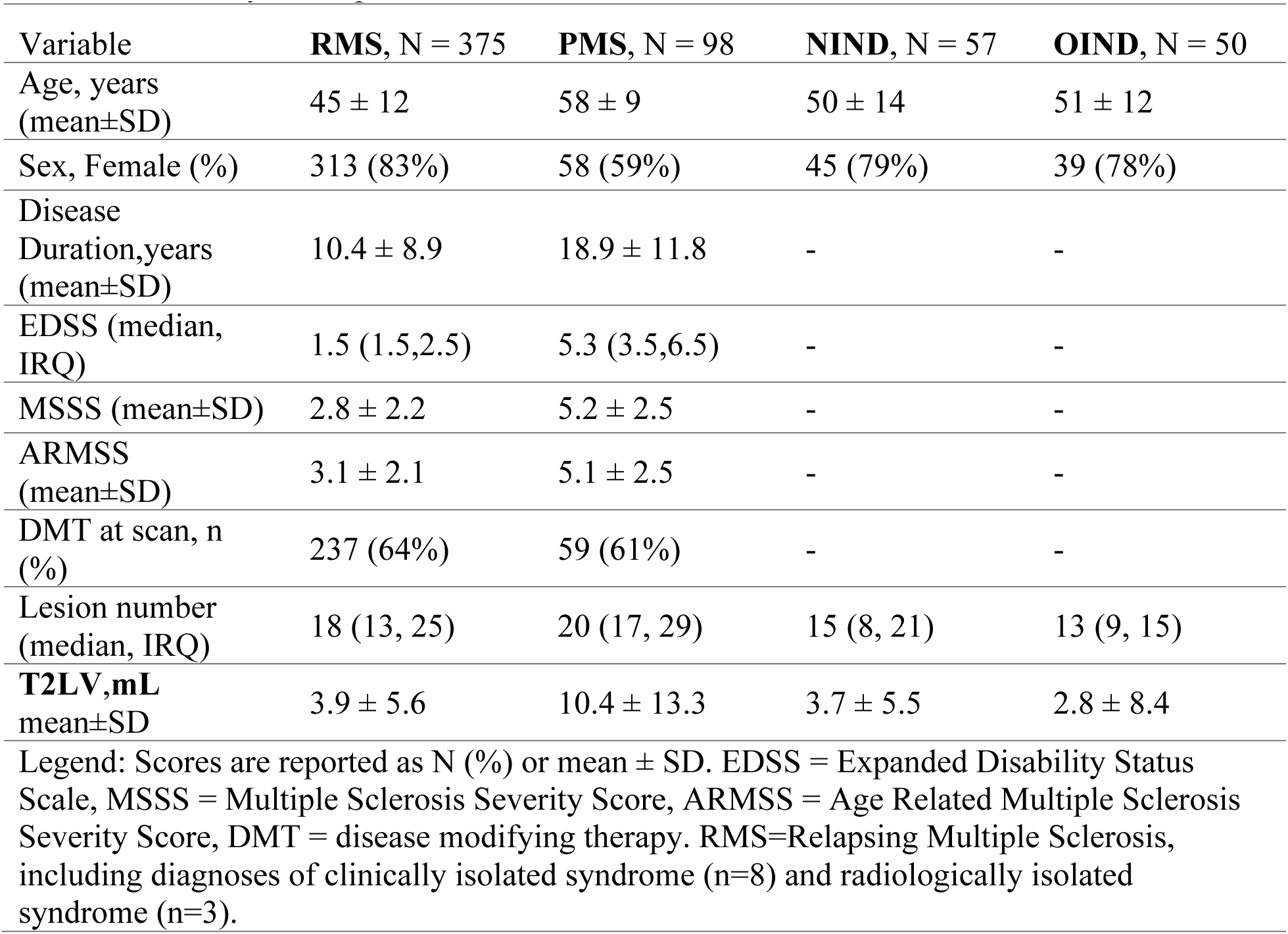
Summary of the patient cohorts

Of patients with NIND and OIND, diagnoses were highly variable. OIND included neurosarcoid (14%), possible MS (14%), unknown (abnormal/inflammatory CSF results and MRI but no identified etiology (12%)), neuro-lupus (12%), NMOSD (6%), neuro-Sjogren’s disease (6%), and others <5% each: MOGAD, primary angiitis of the CNS, CLIPPERS, viral and autoimmune encephalitis, steroid-responsive encephalopathy associated with autoimmune thyroiditis, other rheumatological-associated, other suspected autoimmune-associated, probable CNS lyme, other probable CNS infectious. NIND was comprised of small vessel disease or leukoaraiosis (32%), migraine/headache (20%), neurodegenerative (14%), nonspecific white matter abnormalities in functional neurological disorder or benign disease (9%), and other (25%), a heterogeneous category that included <5% each of various genetic leukodystrophies or epilepsies, toxic-metabolic conditions, fibromyalgia, and idiopathic intracranial hypertension with white matter lesions.

### PRL analysis

Among patients meeting 2017 McDonald criteria for MS (N=473), we observed at least one “definite” PRL in 22% of relapsing patients and 15% of progressive patients. No “definite” PRL were identified in either the NIND or OIND categories, corresponding to a 100% specificity and 21% sensitivity when limited to the category of highest confidence (see **Table 2**). When the confidence threshold was lowered to include “probable” (e.g. “definite” or “probable”), PRL were seen in 37% of RMS and 29% of PMS, as well as 2% (n=1) of both NIND and OIND. This combined category yielded a specificity of 98% and sensitivity of 36%. When reducing confidence to include “possible” lesions (i.e. all categories), 37% of RMS and 56% of PMS patients had at least one lesion meeting these criteria; 16% of NIND and 14% of OIND were also identified as having at least one PRL in this combined category; discrimination specificity was reduced to 85%, and sensitivity was increased to 54%. **Figure 1** exhibits PRL from different confidence categories. One “probable” PRL was identified in both NIND and OIND. The one NIND patient with an identified PRL had been previously treated with glatiramer acetate for suspicion of MS, but this diagnosis was later felt to be unlikely based on further workup including a lack of oligoclonal bands and spinal cord lesions, as well as a history of lacunar strokes and multiple vascular risk factors ultimately felt to be most consistent with (non-inflammatory) small vessel ischemic disease (**Fig 2A**). The diagnosis of the OIND patient was neuro-lupus, although their clinician commented that they suspected co-morbid MS as well (**Fig 2B**).

**Table 2:** Summary of PRL frequencies observed across different diagnostic categories and their associated sensitivity/specificity and interrater agreement

|  |  |  |  |  |  |  |  |
| --- | --- | --- | --- | --- | --- | --- | --- |
| Variable | RMS,<br>N = 375 | PMS,<br>N = 98 | NIND,<br>N = 57 | OIND,<br>N = 50 | Specificity | Sensitivity | Cohen’s<br>κ |
| Definite PRL<br>(≥1) | 82 (22%) | 15 (15%) | 0 (0%) | 0 (0%) | 100% | 21% | 0.91 |
| Probable PRL<br>(≥1) | 103 (27%) | 21 (21%) | 1 (1.8%) | 1 (2.0%) | n/a | n/a | 0.90 |
| Possible PRL<br>(≥1) | 137 (37%) | 41 (42%) | 8 (14%) | 7 (14%) | n/a | n/a | 0.53 |
| Definite or<br>Probable PRL<br>(≥1) | 140 (37%) | 28 (29%) | 1 (1.8%) | 1 (2.0%) | 98% | 36% | 0.94 |
| Definite,<br>Probable, or<br>Possible PRL<br>(≥1) | 201 (54%) | 55 (56%) | 9 (16%) | 7 (14%) | 85% | 54% | 0.74 |

**Caption:** RMS = relapsing-remitting MS (including clinically isolated syndrome, N=8, and radiologically isolated syndrome, N=3); PMS = progressive multiple sclerosis (including primary and secondary progressive forms); NIND = non-inflammatory neurological disease; OIND = other inflammatory neurological disease. n/a = not applicable for calculation, as these categories would naturally be combined with categories of higher confidence in practice.

**Figure 1.**
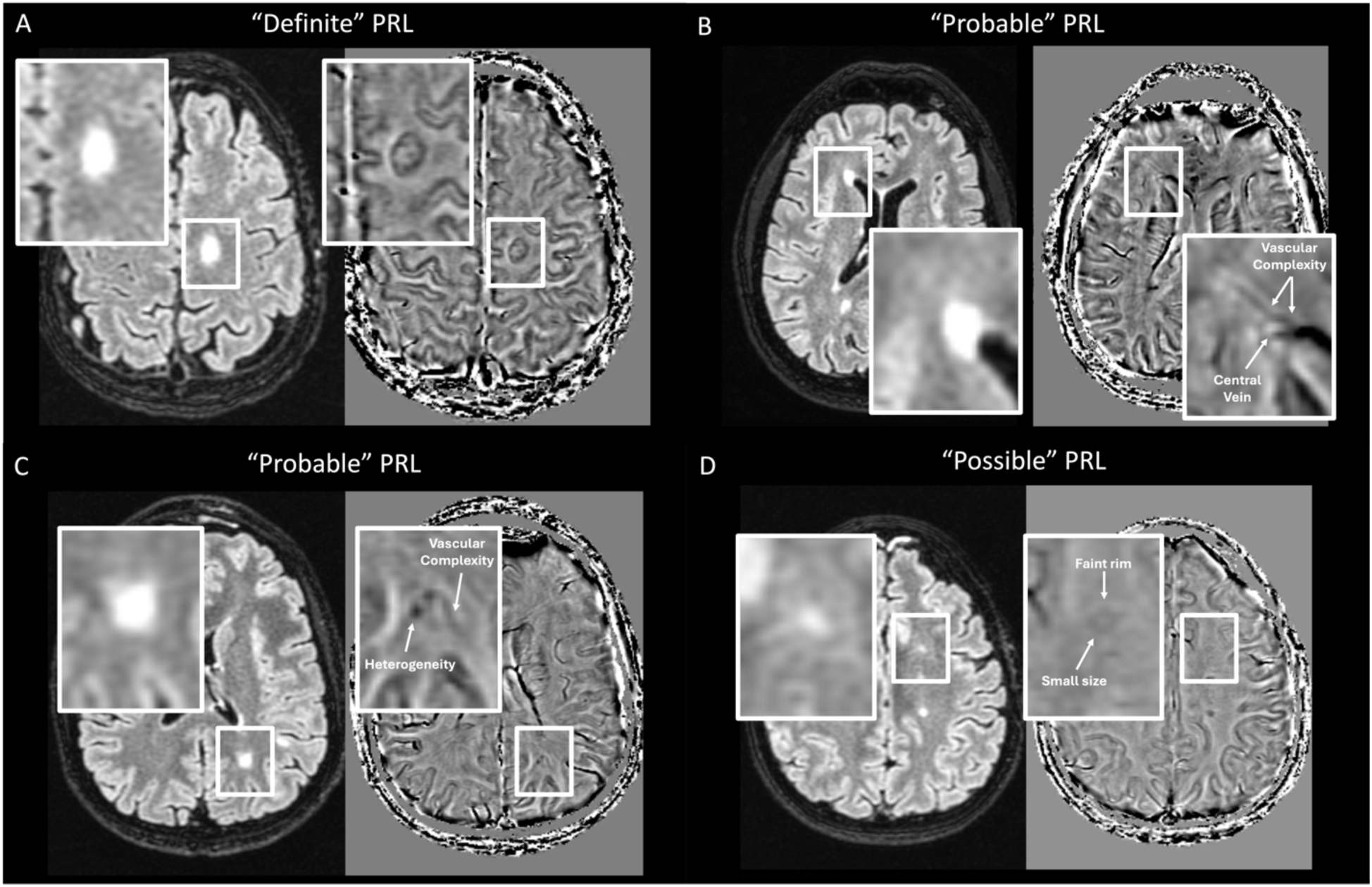
| Examples of PRL ratings of different confidence categories. Axial FLAIR images (left) are coregistered to high-pass filtered phase susceptibility images (SWAN, GE; right) in four different examples demonstrating ratings of (A) “definite”, (B) and (C) “probable”, and (D) “possible” PRL. Small white arrows identify some of the many features that often reduce rater confidence in assessment.

**Figure 2.**
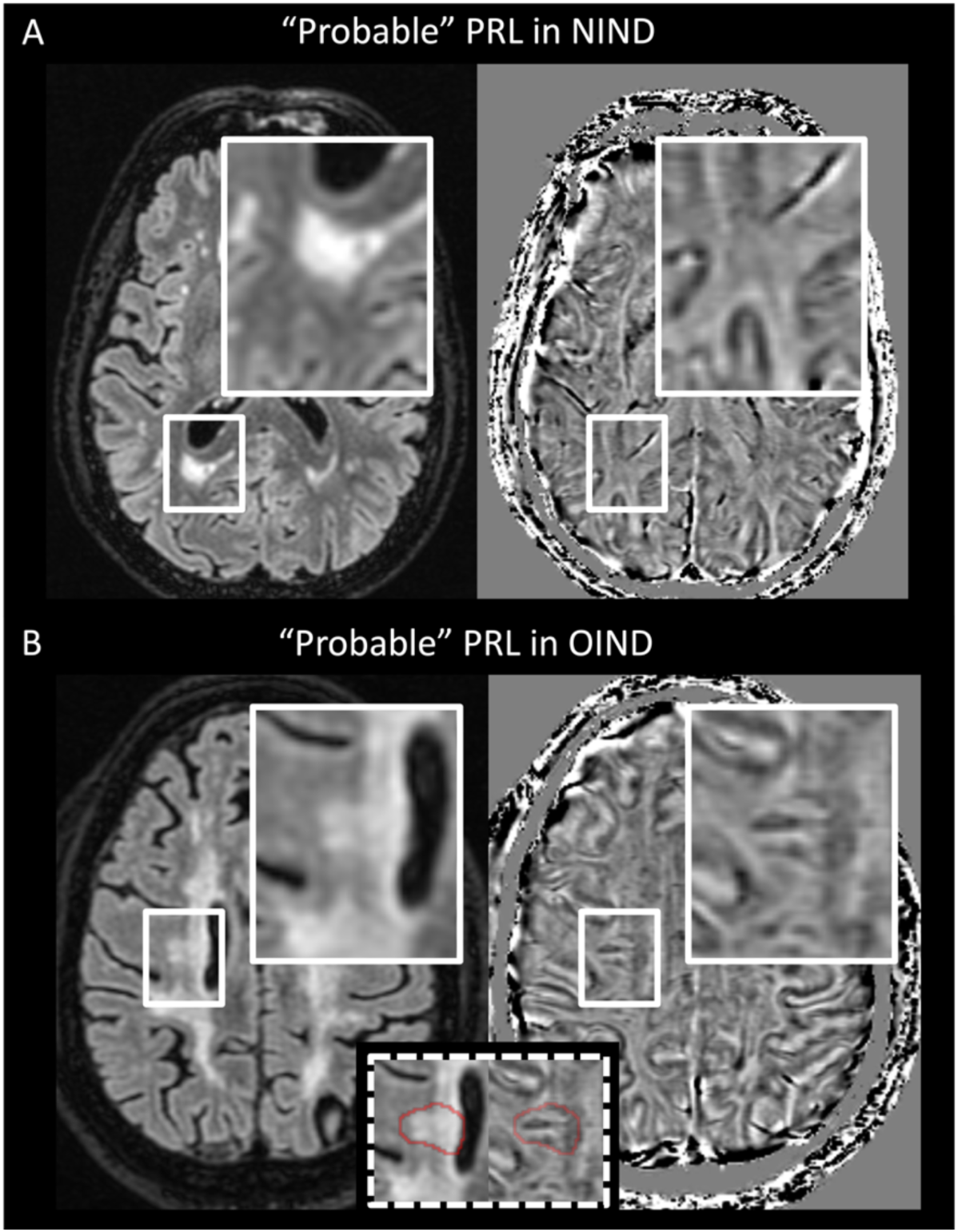
| Probable PRL in non-MS. PRL were rated as “probable” in two patients from the cohort, diagnosed with (A) small vessel disease (non-inflammatory neurological disease) and (B) neurolupus (other inflammatory neurological disease). Axial FLAIR images are on the left and high-pass filtered phase susceptibility images are on the right.

### Interrater analysis

43 patients were evaluated by both raters. At the subject-level, patients having ≥1 “definite” PRL were agreed upon with a κ of 0.91; similarly, patients with ≥1 “probable” PRL showed a κ of 0.90. This agreement was substantially reduced when considering a category of ≥1 “possible” PRL, in which there was a κ = 0.53. Combination categories yielded κ = 0.94 (“definite”/“probable”) and κ = 0.74 (“definite”/“probable”/“possible”). Examples of interrater disagreements can be seen in **Figure 3**.

**Figure 3.**
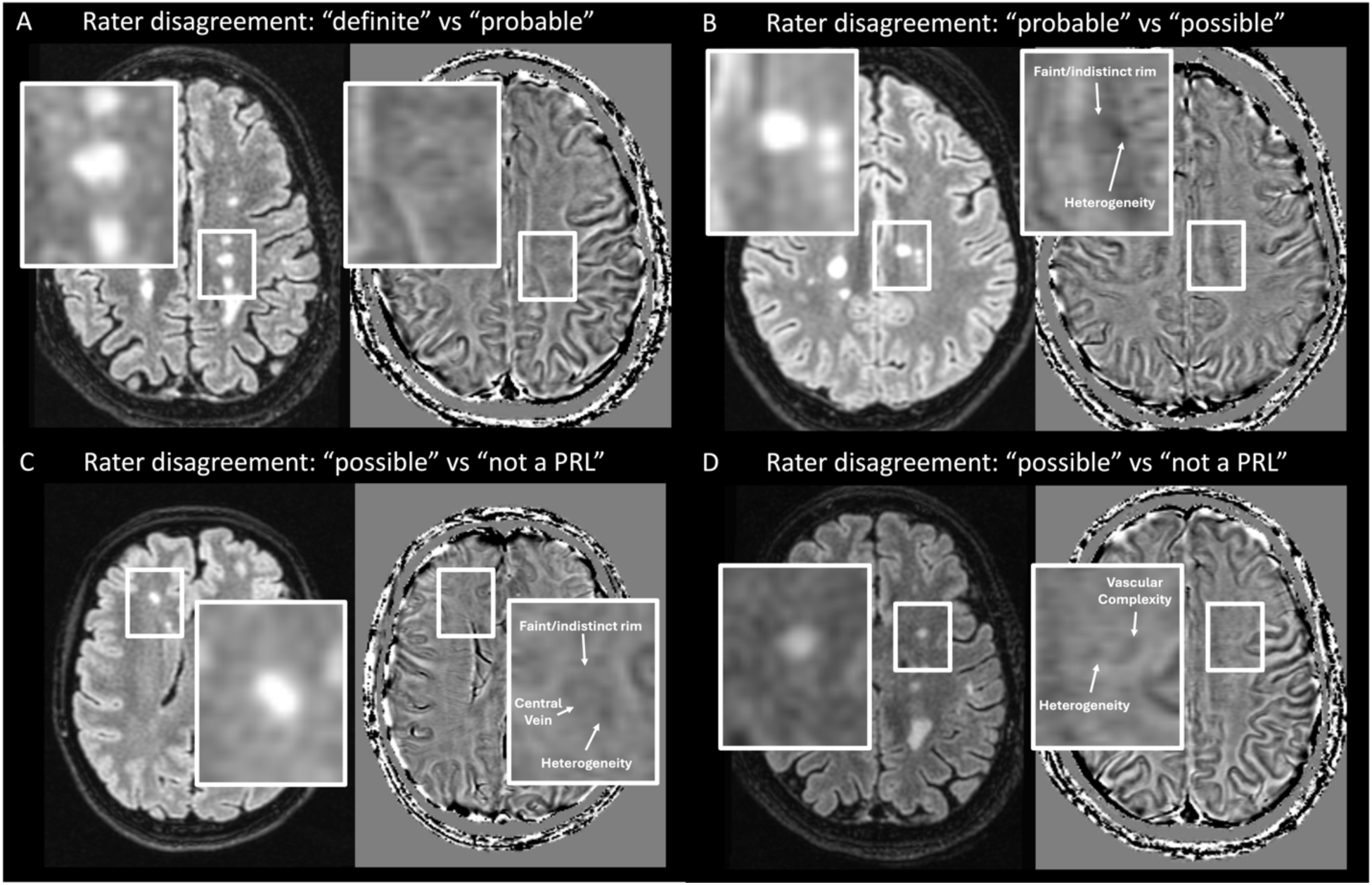
| Examples of rater disagreement at different confidence levels for PRL rating. Frequent sources of disagreement occurred due to faint signal, paramagnetic heterogeneity within the lesion or lesion border, as well as the potential for venous structures to mimic the PRL edge.

### PRL confidence analysis

Raters identified up to 2 of the most significant factors that reduced their perceived confidence in PRL determination. Among the total 563 PRL, raters stated that the following factors played a role: “faint” (33%), “vascular complexity” (25%), “incomplete border” (23%), “heterogeneity” (22%), [being part of a] “confluent [lesion] complex” (17%), “small” (9%), “misalignment” between FLAIR and paramagnetic edges (4%), “artifact” (2%), or “other” (2%). Examples of many of these features can be seen in the **Figures 1**-**3**; **Figure 4** depicts explicit examples of reasons why caution should be advised in lesions that are ultimately very unlikely to represent PRL.

**Figure 4:**
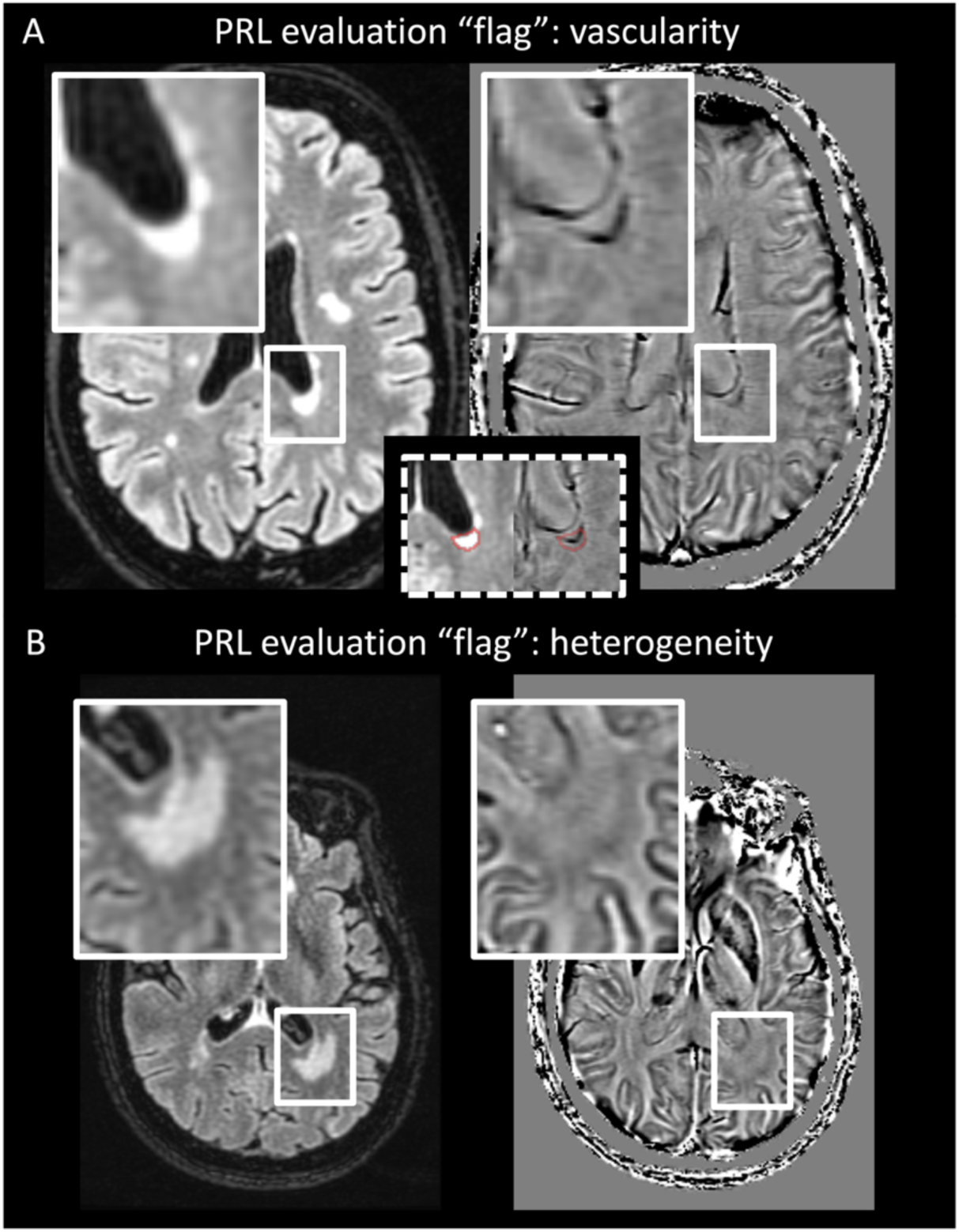
Examples of vascular and heterogeneity “flags” in PRL evaluation that could lead to false positives. (A) demonstrates an enlarged central vein which could readily be mistaken for a paramagnetic rim; an inlay (dotted box) depicts the coregistered edges of the FLAIR lesion which depict an important misalignment. Because the ependyml surface of the ventricle is also paramagnetic, periventricular lesions could be graded with caution. Example (B) shows a faint, thickened, and incomplete paramagnetic signal proximate to the edges of a FLAIR lesion; additional “flags” include dense vascularity traversing the edges. Neither rater identified this as a possible PRL.

### Timed Analysis

30 patients were randomly evaluated. The mean time to identify the first PRL (in cases when “definite” or “probable” PRL ≥1, N=9) was 38±37 seconds (median=22; min=4, max=103). Mean time including cases with “definite” or “probable” PRL=0 (N=21) was 55±31 seconds (median=54; min=4, max=128). Mean time to individually count and summate all PRL, among all included patients was 79±53 seconds (median=66; min=24, max=279).

### Sensitivity Analysis for Early Diagnoses

The most urgent diagnostic need in discriminating MS from its radiological mimics typically occurs within the first months to years of symptom onset. We therefore performed a sensitivity analysis examining only patients with early disease, defined as having had symptom onset within the prior year. All patients from OIND and NIND cohorts were included as the evaluation MRI was typically their index scan. The results of this analysis are presented in Supplementary Tables 1 and 2. This smaller (N=44) cohort had total lesion volumes and numbers in similar proportion to the OIND and NIND comparators, with definite and probable PRL exhibiting the same specificity but with improved sensitivities (definite: 32% vs 21%; definite/probable: 48% vs 36%; definite/probable/possible: 59% vs 54%) due to a higher observed overall frequency of PRL in this early-MS sub-cohort.

## Discussion

PRL are a candidate diagnostic biomarker for MS, given their high demonstrated specificity for MS versus other MRI-mimicking diseases. Here we confirm high discriminative specificity in the largest clinical cohort to date. Strengths of this study include the use of a real-world neuroimmunology clinical cohort and a manufacturer-provided susceptibility-sensitive sequence, as well as the implementation of a novel rater confidence system to enhance clinical translation in the determination of a PRL.

Multiple prior groups have examined sensitivity and specificity of PRL in the determination of MS versus other MRI mimics^3,4,18–25^. In most studies, the specificity of PRL has exceeded 90%. An exception may be Susac disease, in which PRL were commonly observed, although the number of reported cases of this rare disease is small. Unfortunately, our cohort did not include any patients with this diagnosis for comparison, and this remains an area of need for clinical research.

The rationale for implementing a rater confidence system in this study is based on imperfect inter-rater agreement measures and challenges that the raters in this study have previously encountered. This system is congruent with recent consensus criteria determined by the NAIMS Cooperative^6^, but with the additional consideration of a reduced susceptibility signal (i.e. “faint”) as an added factor that may reduce PRL confidence. Our data support the idea — in context of a goal of maximizing diagnostic specificity — that PRL of questionable conviction (“possible”) should be excluded from diagnostic considerations. Raters frequently disagreed on what constituted a “possible” PRL, and although the inclusion of these lesions did improve sensitivity from 36% to 54%, we observed an unacceptably high decrease in specificity, from 98% to 85%. On the other hand, interrater agreement was nearly perfect for the determination of ≥1 PRL of “definite” or “probable” confidence in a given subject, which is reassuring when consideration is given to implementation of PRL as a potential diagnostic biomarker. (Note that our use of “possible” PRL here should not be conflated with the NAIMS consensus^6^ statement’s use of “possible,” which was defined as visualization of PRL in the context of a lack of contrast administration.) And although we provide a suggested PRL evaluation outline here, we nonetheless acknowledge that this “confidence categorization” assessment is ultimately qualitative in nature, remaining untested in wider neuroradiological practice.

This study was designed with clinical translation in mind. PRL evaluations were rapid (55 seconds), especially when only considering the time-to-first PRL identification. Sensitivity could likely be improved using a higher in-plane resolution, at the expense of longer scan times and consequent increase in risk of motion artifacts. Determining an “optimized” resolution and protocol to detect PRL with each manufacturer remains and area of high clinical need. For example, the use of a single GE “SWAN” sequence here may not directly compare to other manufacture sequences (such as Philips “SWIp” or Siemens “SWI”), although we note from the literature as well as our own experience that PRL can been readily visualized using these manufacturer sequences as well. We also implemented a single rater system for lesion assessment, as would be reflected in practice. Scans were coregistered as part of our preprocessing, an important step that is included in many modern PACS software and does not often represent a translational limitation in neuroradiology practice. We did not use the T1 sequence in PRL determination, which may aid in cases of poor FLAIR-susceptibility colocalization (here affecting 4% of PRL assessments). Although raters were allowed to view the lower-resolution multiplanar reformats here, this occurred only rarely as the 3mm out-of-plane thickness added little perceived confidence, and slowed the evaluations. Acquiring thinner slice thicknesses would facilitate orthogonal evaluations, but also may come at the expense of increased scan times depending on the acquisition technique. Notably, much of the early research with PRL relied on a high-resolution, isotropic, segmented-EPI acquisition (optimized for high contrast between lesions and central veins^26^) in which orthogonal reformats provided more detailed information; this sequence is unfortunately not yet widely available as a manufacturer product sequence.

Interrater agreement was almost perfect for “probable” and “definite” PRL, although the raters here had worked together previously and generally formed a “local” consensus for what constitutes a PRL. Although this could have artificially inflated agreement, we note that raters nonetheless continued to disagree on what constitutes a “possible” PRL. Additional research is needed to evaluate and optimize the rater training process.

Last, care should be made to find an appropriate contrast and window for viewing PRL. In our experience with GE (and Philips) susceptibility protocols, PRL histogram data are normally distributed around 0 (with a range that should fall between *π* and negative *π,* sometimes with a scaling factor), and we suggest a window covering approximately ±1.4 SD around the mean of the data to obtain good PRL contrast. However, we did not systematically explore this issue, which is an important topic for future research directions.

## Conclusions

The observation of at least one PRL of “probable” or “definite” confidence is associated with very high discriminative specificity for MS versus other common MRI mimics. Raters should remain conservative in their PRL assessments for diagnostic purposes and be familiar with the many features that degrade confidence in lesion assessments. PRL are readily and rapidly visualized using a manufacturer protocol in the clinical routine. These data, using a large cohort of real-world neuroimmunology cases, support proposals for PRL integration into future diagnostic MS criteria.

## Data Availability

Deidentified data are available to qualified collaborators, following a signed institutional data sharing agreement.

## Acknowledgements

CCH receives extramural research funding from NIH K23NS126718. DSR is funded by the Intramural Research Program of NINDS. The content is solely the responsibility of the authors and does not necessarily represent the official views of the National Institutes of Health. We thank colleagues including Drs. Roberto Bomprezzi, Idanis Berrios-Morales and Raffaella Umeton as well as Celia Gomes-McGillivray, Nimmy Francis, Aurelie Martin-Puig, and Mariana Kurban at the University of Massachusetts for contributions to patient recruitment in ongoing, longitudinal observational studies. We thank the family of Mrs. Jane Gagne, in memory of Edward Gagne, for their ongoing research support.

## Author contributions

CCH: conceptualization, methodology, software, formal analysis, investigation, resources, writing-original draft, writing-review and editing, visualization, funding acquisition SD: methodology, investigation, writing-review and editing MD: resources, investigation, writing-review and editing JB: methodology, supervision, writing-review and editing RB: resources, supervision, writing-review and editing CI: resources, supervision, investigation, writing-review and editing DSR: conceptualization, methodology, supervision, writing-original draft, writing-review and editing

