## Supplemental Table for "Paramagnetic Rim Lesions are Highly Specific for Multiple Sclerosis in Real-World Data"

| **Supplementary Table 1: Sensitivity analysis cohort of patients within the first year of symptom onset** | | | | |
| --- | --- | --- | --- | --- |
| **Variable** | **RMS, N = 43** | **PMS, N = 1** | **NIND, N = 57** | **OIND, N = 50** |
| Age, years (mean±SD) | **39 ± 10** | **56** | **50 ± 14** | **51 ± 12** |
| Sex, Female (%) | **31 (72%)** | **1 (100%)** | **45 (79%)** | **39 (78%)** |
| Disease Duration, years (mean±SD) | **0.4 ± 0.6** | **0.8** | **-** | **-** |
| EDSS (median, IRQ) | **1.5 (1.0,2.0)** | **5.0** | **-** | **-** |
| MSSS (mean±SD) | **4.1 ± 1.6** | **9.5** | **-** | **-** |
| ARMSS (mean±SD) | **3.0 ± 1.4** | **5.3** | **-** | **-** |
| DMT at scan, n (%) | **15 (36%)** | **1 (100%)** | **-** | **-** |
| Lesion number (median, IRQ) | **14 (12,18)** | **20** | **15 (8,21)** | **13 (9,15)** |
| T2LV,mL mean±SD | **2.1 ± 2.0** | **2.0** | **3.7 ± 5.5** | **2.8 ± 8.4** |
| Legend: Scores are reported as N (%) or mean ± SD. EDSS = Expanded Disability Status Scale, MSSS = Multiple Sclerosis Severity Score, ARMSS = Age Related Multiple Sclerosis Severity Score, DMT = disease modifying therapy; RMS=Relapsing Multiple Sclerosis, including diagnoses of clinically isolated syndrome (n=5). | | | | |

| Supplementary Table 2: Summary of PRL frequencies and sensitivity/specificity, observed across different diagnostic categories in the first year following symptom onset | | | | | | |
| --- | --- | --- | --- | --- | --- | --- |
| Variable | **RMS,**  **N = 43** | **PMS,**  **N = 1** | **NIND,**  **N = 57** | **OIND,**  **N = 50** | **Specificity** | **Sensitivity** |
| Definite PRL (≥1) | 14 (33%) | 0 (0%) | 0 (0%) | 0 (0%) | 100% | 32% |
| Probable PRL (≥1) | 14 (33%) | 1 (100%) | 1 (1.8%) | 1 (2.0%) | n/a | n/a |
| Possible PRL (≥1) | 13 (30%) | 1 (100%) | 8 (14%) | 7 (14%) | n/a | n/a |
| Definite or Probable PRL (≥1) | 20 (47%) | 1 (100%) | 1 (1.8%) | 1 (2.0%) | 98% | 48% |
| Definite, Probable, or Possible PRL (≥1) | 25 (58%) | 1 (100%) | 9 (16%) | 7 (14%) | 85% | 59% |
| Caption: RMS = relapsing-remitting MS (including clinically isolated syndrome, N=8, and radiologically isolated syndrome, N=3); PMS = progressive multiple sclerosis (including primary and secondary progressive forms); NIND = non-inflammatory neurological disease; OIND = other inflammatory neurological disease. n/a = not applicable for calculation, as these categories would naturally be combined with categories of higher confidence in practice. | | | | | | |
